# Gut microbiota may underlie the predisposition of healthy individuals to COVID-19

**DOI:** 10.1101/2020.04.22.20076091

**Authors:** Wanglong Gou, Yuanqing Fu, Liang Yue, Geng-dong Chen, Xue Cai, Menglei Shuai, Fengzhe Xu, Xiao Yi, Hao Chen, Yi Zhu, Mian-li Xiao, Zengliang Jiang, Zelei Miao, Congmei Xiao, Bo Shen, Xiaomai Wu, Haihong Zhao, Wenhua Ling, Jun Wang, Yu-ming Chen, Tiannan Guo, Ju-Sheng Zheng

## Abstract

The COVID-19 pandemic is spreading globally with high disparity in the susceptibility of the disease severity. Identification of the key underlying factors for this disparity is highly warranted. Here we describe constructing a proteomic risk score based on 20 blood proteomic biomarkers which predict the progression to severe COVID-19. We demonstrate that in our own cohort of 990 individuals without infection, this proteomic risk score is positively associated with proinflammatory cytokines mainly among older, but not younger, individuals. We further discovered that a core set of gut microbiota could accurately predict the above proteomic biomarkers among 301 individuals using a machine learning model, and that these gut microbiota features are highly correlated with proinflammatory cytokines in another set of 366 individuals. Fecal metabolomic analysis suggested potential amino acid-related pathways linking gut microbiota to inflammation. This study suggests that gut microbiota may underlie the predisposition of normal individuals to severe COVID-19.

## Introduction

With the coronavirus disease 2019 (COVID-19) defined as ‘global pandemic’ and spreading worldwide at an unprecedented speed, more than two million individuals have been infected globally since its first detection in December 2019 to mid-April 2020 (WHO, 2020). So far, many research papers have been published to characterize the clinical features of the COVID-19 patients, revealing that those individuals who are older, male or having other clinical comorbidities are more likely to develop into severe COVID-19 cases (Chen et al., 2020; Huang et al., 2020). Yet, little is known about the potential biological mechanisms or predictors for the susceptibility of the disease.

It is known that COVID-19 is caused by severe acute respiratory syndrome coronavirus 2 (SARS-CoV-2), which enters human cells by binding to angiotensin converting enzyme 2 (ACE2) as its receptor (Yan et al., 2020). Of note, ACE2 is an important regulator of intestinal inflammation, and that the expression of ACE2 is higher in the ileum and colon than in lung (Hashimoto et al., 2012; Zhang et al., 2020). ACE2 also has a major impact on the composition of gut microbiota, thus affecting cardiopulmonary diseases (Cole-Jeffrey et al., 2015). Moreover, over 60% of patients with COVID-19 report evidence of gastrointestinal symptoms, such as diarrhoea, nausea and vomiting, and that patients with gastrointestinal symptoms had overall more severe/critical diseases (Jin et al., 2020; Lin et al., 2020; Ng and Tilg, 2020). Taken together, the available evidence suggests a potential role of gut microbiota in the susceptibility of COVID-19 progression and severity.

Based on a recent investigation into the blood biomarkers of COVID-19 patients, we identified a set of proteomic biomarkers which could help predict the progression to severe COVID-19 among infected patients (Shen et al., 2020). The newly discovered proteomic biomarkers may help early prediction of severe COVID-19. However, the question remains as to whether this set of proteomic biomarkers could be used in healthy (non-infected) individuals to help explain the disease susceptibility. It is also unclear whether gut microbiota could regulate these blood proteomic biomarkers among healthy individuals.

To address the above unresolved questions, we integrated blood proteomics data from 31 COVID-19 patients and multi-omics data from a Chinese population without infection living in Guangzhou, involving 2413 participants (Figure 1; Figure S1; Table S1). Based on the COVID-19 patient data, we constructed a blood proteomic risk score (PRS) for the prediction of COVID-19 progression to clinically severe phase. Then, among 990 healthy individuals with the data of proteome and blood inflammatory biomarkers, we investigated the association of the COVID-19-related PRS with inflammatory biomarkers as a verification of the PRS with disease susceptibility in normal non-infected individuals. Next, we identified core gut microbiota features which predicted the blood proteomic biomarkers of COVID-19 using a machine-learning model. We conducted further fecal metabolomics analysis to reveal potential biological mechanisms linking gut microbiota to the COVID-19 susceptibility among non-infected individuals. Finally, we demonstrated the contribution of 40 host and environmental factors to the variance of the above identified core gut microbiota features.

**Figure 1.**
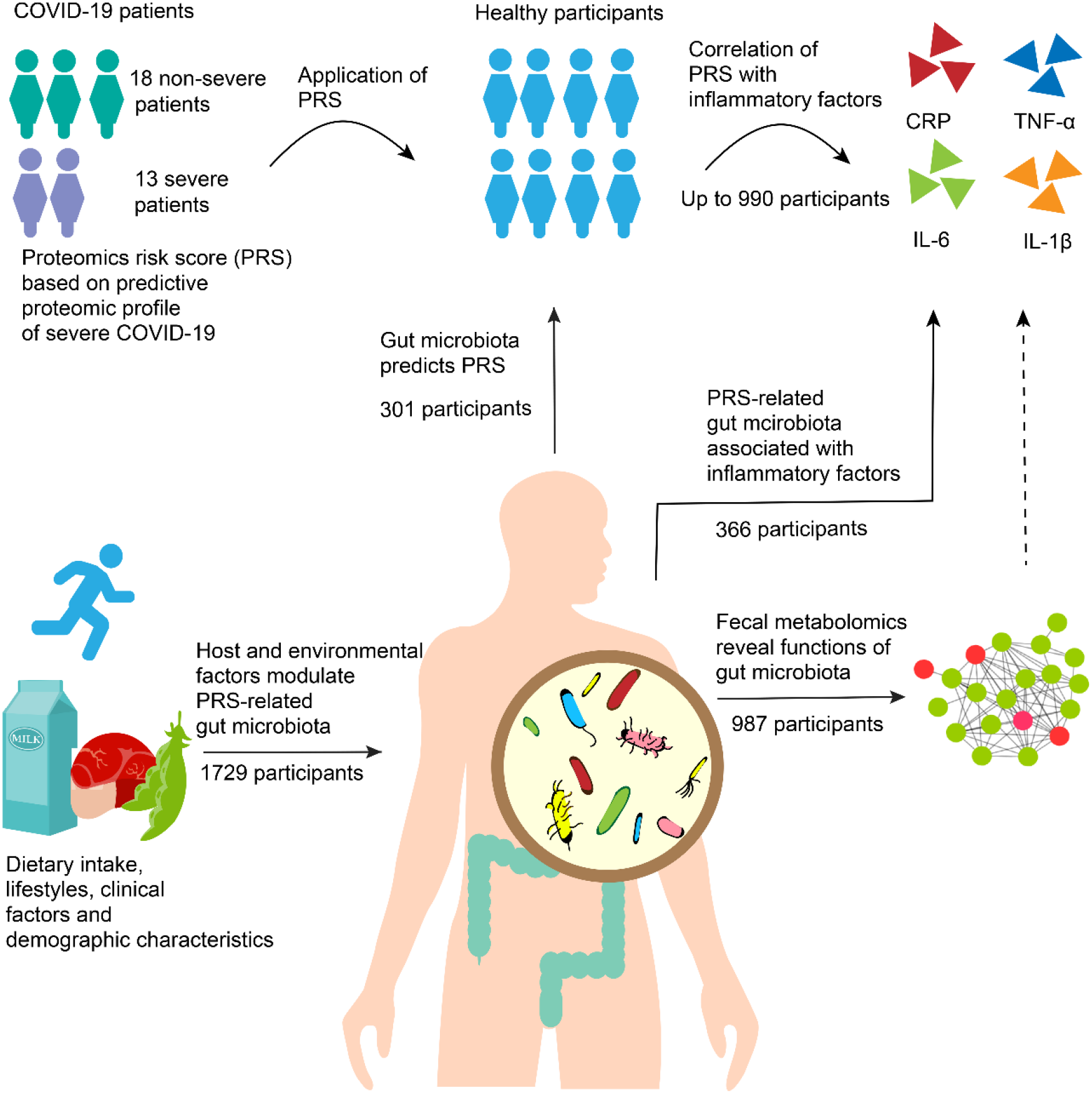
Study design and analysis pipeline. Study overview. 1) constructing a novel COVID-19 blood proteomic risk score (PRS) among 31 COVID-19 patients (18 non-severe cases and 13 severe cases). 2) Applicating the PRS in healthy participants, and further linking it to host inflammatory status (n=990). 3) Investigating the potential role of gut microbiota in predicting the PRS of COVID-19 based on a machine-learning method (n=301). 4) Assessing the relationships between the PRS-related gut microbiota and inflammatory factors (n=336). 5) Fecal metabolomics analysis reveals function of gut microbiota on host metabolism (n=987). 6) Investigating the impact of host and environmental factors on PRS-related core microbial OTUs (n=1729).

## Results

### Predictive proteomic profile for severe COVID-19 is correlated with inflammatory factors among healthy individuals

Based on a prior serum proteomic profiling of COVID-19 patients, 22 proteomic biomarkers contributed to the prediction of progression to severe COVID-19 status (Shen et al., 2020).Using this cohort, we constructed a blood PRS among the 31 COVID-19 patients (18 non-severe cases and 13 severe cases) based on 20 proteomic biomarkers (Table S2). We only used 20 of the 22 proteins for our PRS construction because 2 proteins were unavailable in our large proteomics database among non-infected participants for the further analysis. Among the COVID-19 patients, Poisson regression analysis indicated that per 10% increment in the PRS there was associated a 57% higher risk of progressing to clinically severe phase (RR, 1.57; 95% CI, 1.35-1.82; Figure 2A), in support of the PRS as being a valid proxy for the predictive biomarkers of severe COVID-19.

**Figure 2.**
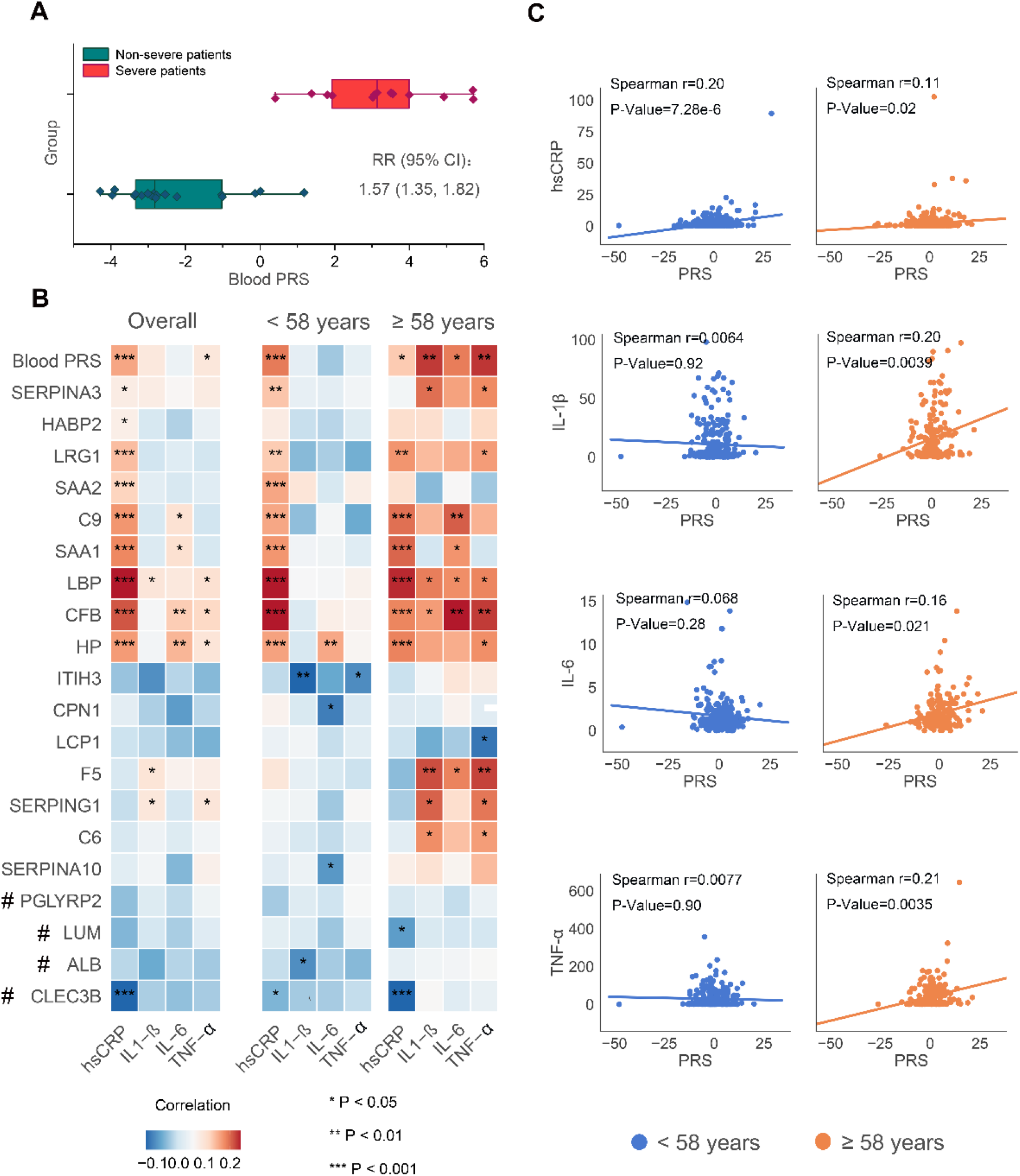
Predictive proteomic profile for severe COVID-19 is correlated with pro-inflammatory factors among healthy individuals. **(A)** The associations of COVID-19-related blood proteomic biomarkers and proteomic risk score (PRS) with host inflammatory markers. 990 participants were involved in this analysis. # protein down-regulated in severe patients, else, up-regulated. **(B)** The correlation of the above blood proteomic biomarkers and PRS with host inflammatory markers stratified by the median age of participants (<58 years or ≥58 years). The color of the heatmap indicates the Spearman correlation coefficients (blue-negative, red-positive). **(C)** The correlation of the PRS with individual host inflammatory markers stratified by the median age of participants (<58 years or ≥58 years).

To explore the potential implication of the PRS among non-COVID-19 individuals, we constructed the PRS using the same set of 20 blood proteins among a cohort of non-infected participants with data of both proteomics and inflammatory markers (n=990). The blood proteomic data was based on the baseline serum samples of the cohort (Figure S1). We investigated the correlation between the PRS and blood inflammatory markers IL-1β, IL-6, TNF-α and hsCRP. The PRS had a significantly positive correlation with serum concentrations of hsCRP and TNF-α (p<0.001 and p<0.05, respectively), but not other markers (Figure 2B). As age and sex are very important factors related to the susceptibility to SARS-CoV-2 infection, we performed subgroup analysis stratified by age (<58 years vs. ≥58 years, with 58 years as the median age of this cohort) and sex. Interestingly, we found that higher PRS was significantly correlated with higher serum concentrations of all the aforementioned inflammatory markers among older individuals (>58 years, n=493), but not among younger individuals (≤58 years, n=497) (Figure 2B and 2C). The PRS did not show any differential association with the inflammatory markers by sex (Figure S2). Whether the identified proteomic changes causally induce immune activation or consequences of the immune response are not clear at present, but the finding supports the hypothesis that the PRS may act as a biomarker of unbalanced host immune system, especially among older adults.

### Core microbiota features predict COVID-19 proteomic risk score and host inflammation

To investigate the potential role of gut microbiota in the susceptibility of healthy individuals to COVID-19, we next explored the relationship between the gut microbiota and the above COVID-19-related PRS in a sub-cohort of 301 participants with measurement of both gut microbiota (16s rRNA) and blood proteomics data (Figure S1). Gut microbiota data were collected and measured during a follow-up visit of the cohort participants, with a cross-sectional subset of the individuals (n=132) having blood proteomic data at the same time point as the stool collection and another independent prospective subset of the individuals (n=169) having proteomic data at a next follow-up visit ∼3 years later than the stool collection.

Among the cross-sectional subset, using a machine learning-based method: LightGBM and a very conservative and strict tenfold cross-validation strategy, we identified 20 top predictive operational taxonomic units (OTUs), and this subset of core OTUs explained an average 21.5% of the PRS variation (mean out-of-sample R^2^=0.215 across ten cross-validations). The list of these core OTUs along with their taxonomic classification is provided in Table S3. These OTUs were mainly assigned to *Bacteroides* genus, *Streptococcus* genus, *Lactobacillus* genus, *Ruminococcaceae* family, *Lachnospiraceae* family and *Clostridiales* order.

To test the verification of the core OTUs, the Pearson correlation analysis showed the coefficient between the core OTUs-predicted PRS and actual PRS reached 0.59 (p<0.001), substantially outperforming the predictive capacity of other demographic characteristics and laboratory tests including age, BMI, sex, blood pressure and blood lipids (Pearson’s r =0.154, p=0.087) (Figure 3A). Additionally, we used co-inertia analysis (CIA) to further test co-variance between the 20 identified core OTUs and 20 predictive proteomic biomarkers of severe COVID-19, outputting a RV coefficient (ranged from 0 to 1) to quantify the closeness. The results indicated a close association of these OTUs with the proteomic biomarkers (RV=0.12, p<0.05) (Figure S3A). When replicating this analysis stratified by age, significant association was observed only among older participants (age≥58, n=66; RV=0.22, p<0.05) (Figure S3B and S3C).

**Figure 3.**
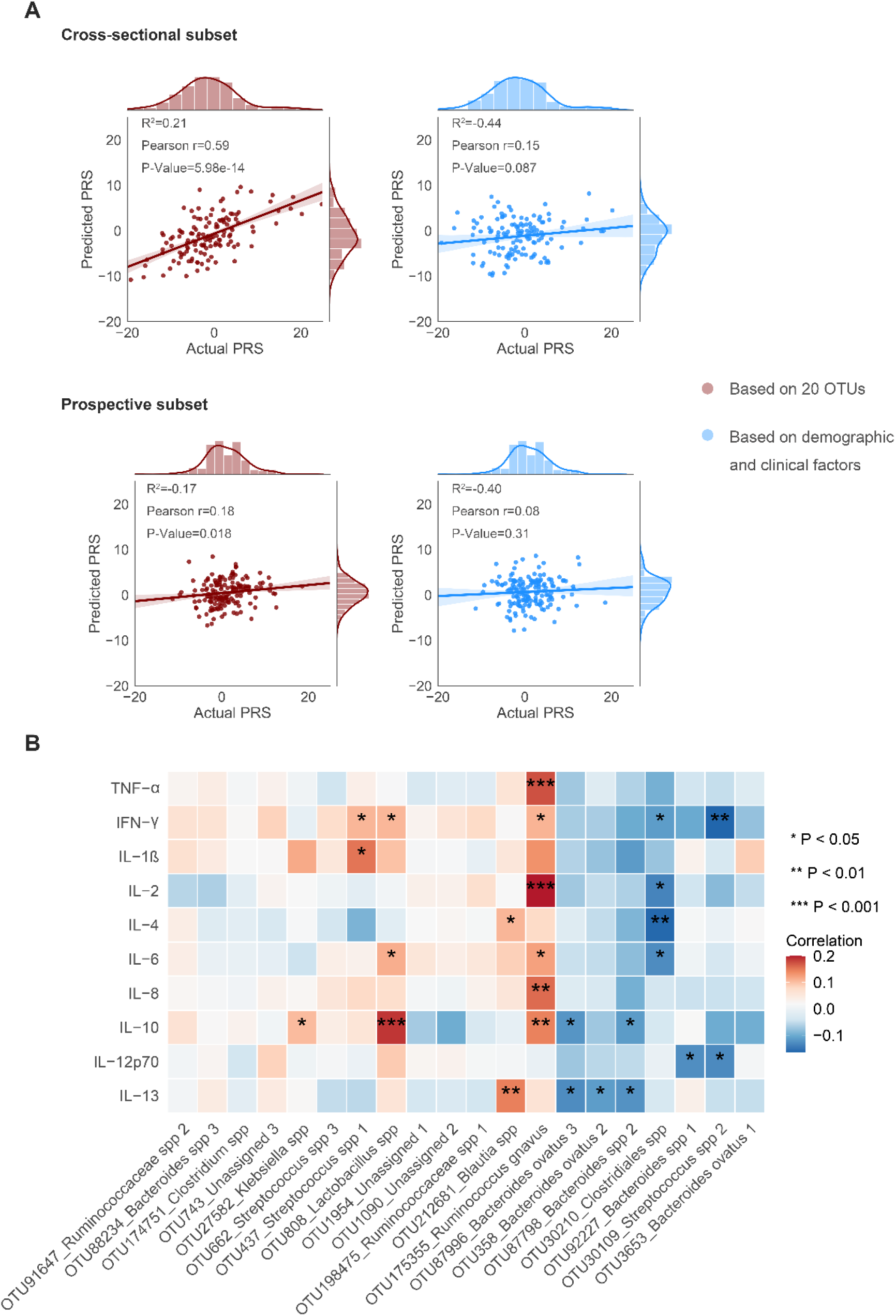
Core microbiota features predict COVID-19 proteomic risk score (PRS) and host inflammation. **(A)** Plots of out-of-sample predicted PRS versus actual PRS based on top 20 ranked OTUs or demographic/clinical factors (age, sex BMI, fasting glucose, HDL, LDL, TC, TG, DBP, and SBP) using LightGBM with 10-fold cross-validation. The plots in the first row indicate the model performance among cross-sectional subset of individuals (n=132); the plots in the second row indicate the model performance among prospective subset of individuals (n=169).The mean R^2^ across the 10 cross validations, Pearson r of predicted values versus actual values, and corresponding P-value are shown in the figures. **(B)** The correlation of the core microbial OTUs and host inflammatory cytokines (n=336). The color of the heatmap indicates the Spearman correlation coefficients (blue-negative, red-positive).

Importantly, the above results from cross-sectional analyses were successfully replicated in the independent prospective subset of 169 individuals, which showed a Pearson’s r of 0.18 between the core OTUs-predicted PRS versus actual PRS (p<0.05), also outperforming the predictive capacity of the above demographic characteristics and laboratory tests (Pearson’s r =0.08, p=0.31) (Figure 3A). These findings support that change in the gut microbiota may precede the change in the blood proteomic biomarkers, inferring a potential causal relationship.

To further verify the reliability of these core OTUs, in another larger independent sub-cohort of 366 participants (Figure S1), we examined the cross-sectional relationship between the core OTUs and 10 host inflammatory cytokines including IL-1β, IL-2, IL-4, IL-6, IL-8, IL-10, IL-12p70, IL-13, TNF-α and IFN-γ, and found 11 microbial OTUs were significantly associated with the inflammatory cytokines (Figure 3B). Specifically, *Bacteroides* genus, *Streptococcus* genus and *Clostridiales* order were negatively correlated with most of the tested inflammatory cytokines, whereas *Ruminococcus* genus, *Blautia* genus and *Lactobacillus* genus showed positive associations.

### Fecal metabolome may be the key to link the PRS-related core microbial features and host inflammation

We hypothesized that the influences of the core microbial features on the PRS and host inflammation were driven by some specific microbial metabolites. So we assessed the relationship between the core gut microbiota and fecal metabolome among 987 participants, whose fecal metabolomics and 16s rRNA microbiome data were collected and measured at the same time point during the follow-up visit of the participants (Figure S1). After correction for the multiple testing (FDR<0.05), a total of 183 fecal metabolites had significant correlations with at least one selected microbial OTU. Notably, 45 fecal metabolites, mainly within the categories of amino acids, fatty acids and bile acids, showed significant associations with more than half of the selected microbial OTUs (Figure 4A), these metabolites might play a key role in mediating the effect of the core gut microbiota on host metabolism and inflammation.

**Figure 4.**
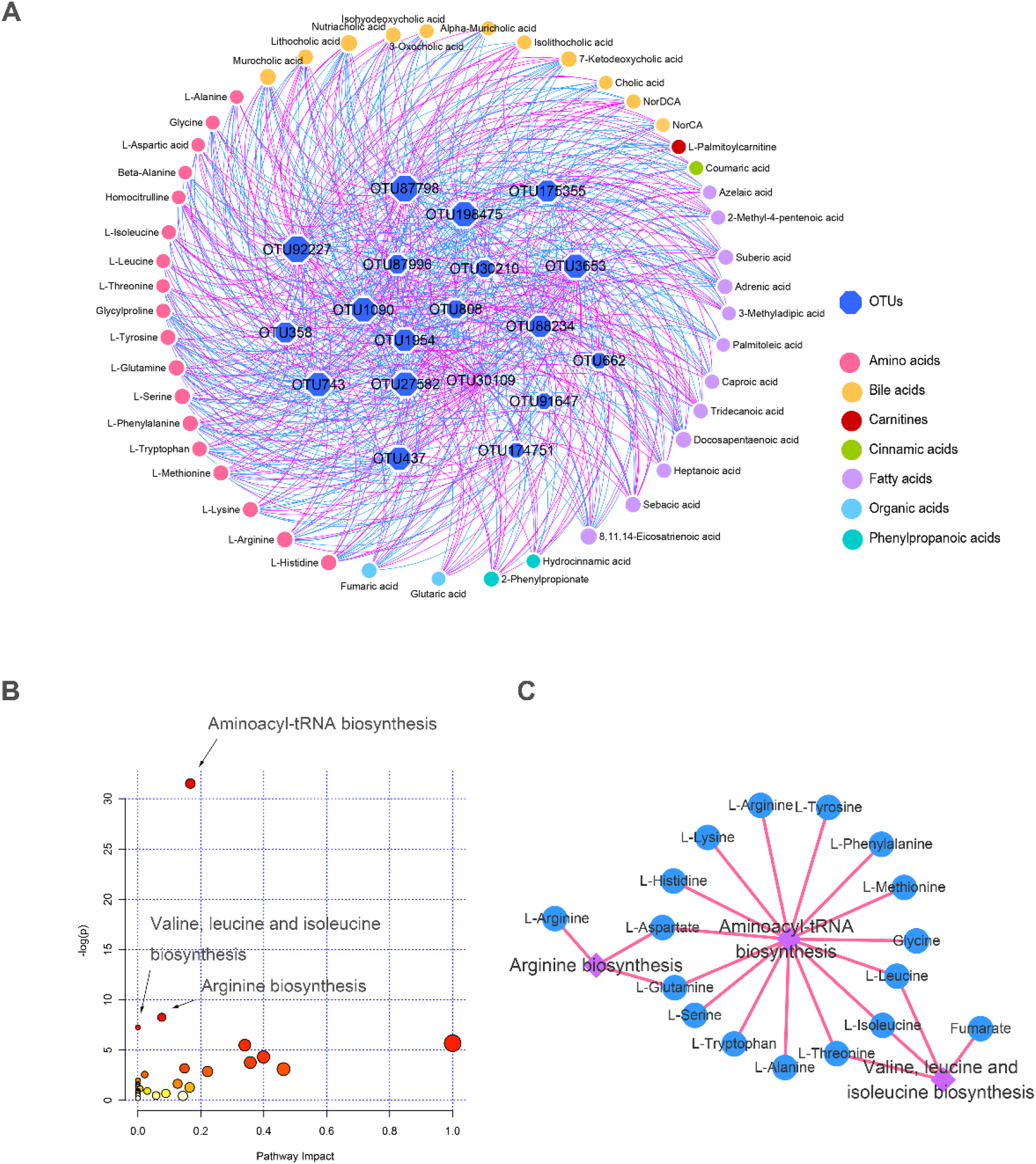
Fecal metabolome may be the key to link the proteomic risk score-related core microbial features and host inflammation. **(A)** Associations of the core microbial OTUs with fecal metabolites (n=987). The relationships between microbial OTUs and fecal metabolites was assessed by a linear regression model adjusting for age, sex, BMI. Multiple testing was adjusted using Benjamini and Hochberg method, with a false discovery rate (FDR) of <0.05 being considered statistically significant. We only presented metabolites showing significant associations with more than half of the core microbial OTUs (n=20) in the figure. Sizes of the nodes represent the number of OTUs related with fecal metabolites. Red edge, β-coefficient >0; blue edge, β-coefficient <0. **(B)** Pathway analysis for the core fecal metabolites (shown in part A) using MetaboAnalyst 4.0 (Chong et al., 2019). **(C)** Metabolites enriched in the significant pathways (shown in part B).

Based on these key metabolites, we performed metabolic pathway analysis to elucidate possible biological mechanisms. The results showed that these 45 fecal metabolites were mainly enriched in three pathways, namely aminoacyl-tRNA biosynthesis pathway, arginine biosynthesis pathway, and valine, leucine and isoleucine biosynthesis pathway (Figure 4B). There were 15 fecal metabolites involved in the aminoacyl-tRNA biosynthesis pathway, which is responsible for adding amino acid to nascent peptide chains and is a target for inhibiting cytokine stimulated inflammation (Figure 4C). Additionally, 4 metabolites were associated with arginine biosynthesis pathway and 3 metabolites were enriched in valine, leucine and isoleucine (known as branch-chain amino acids, BCAAs) biosynthesis pathway (Figure 4C).

### Host and environmental factors modulate the PRS-related core microbial OTUs

As demographic, socioeconomic, dietary and lifestyle factors may all be closely related to the gut microbiota, we explored the variance contribution of these host and environmental factors for the identified core OTU composition. A total of 40 items belonging to two categories (i.e., demographic/clinical factors and dietary/nutritional factors) were tested (Figure 5), which together explained 3.6% of the variation in interindividual distance of the core OTU composition (Bray-Curtis distance). In the demographic/clinical factors which explained 2.4% of the variation, we observed associations of 9 items (i.e., sex, education, physical activity, diastolic blood pressure, blood glucose, blood lipids and medicine use for type 2 diabetes) with inter-individual distances in the core OTU composition (PERMANOVA, p<0.05; Figure 5). While in the dietary/nutritional category (1.1% variance was explained), only dairy consumption significantly contributed to the variance of the core OTU composition.

**Figure 5.**
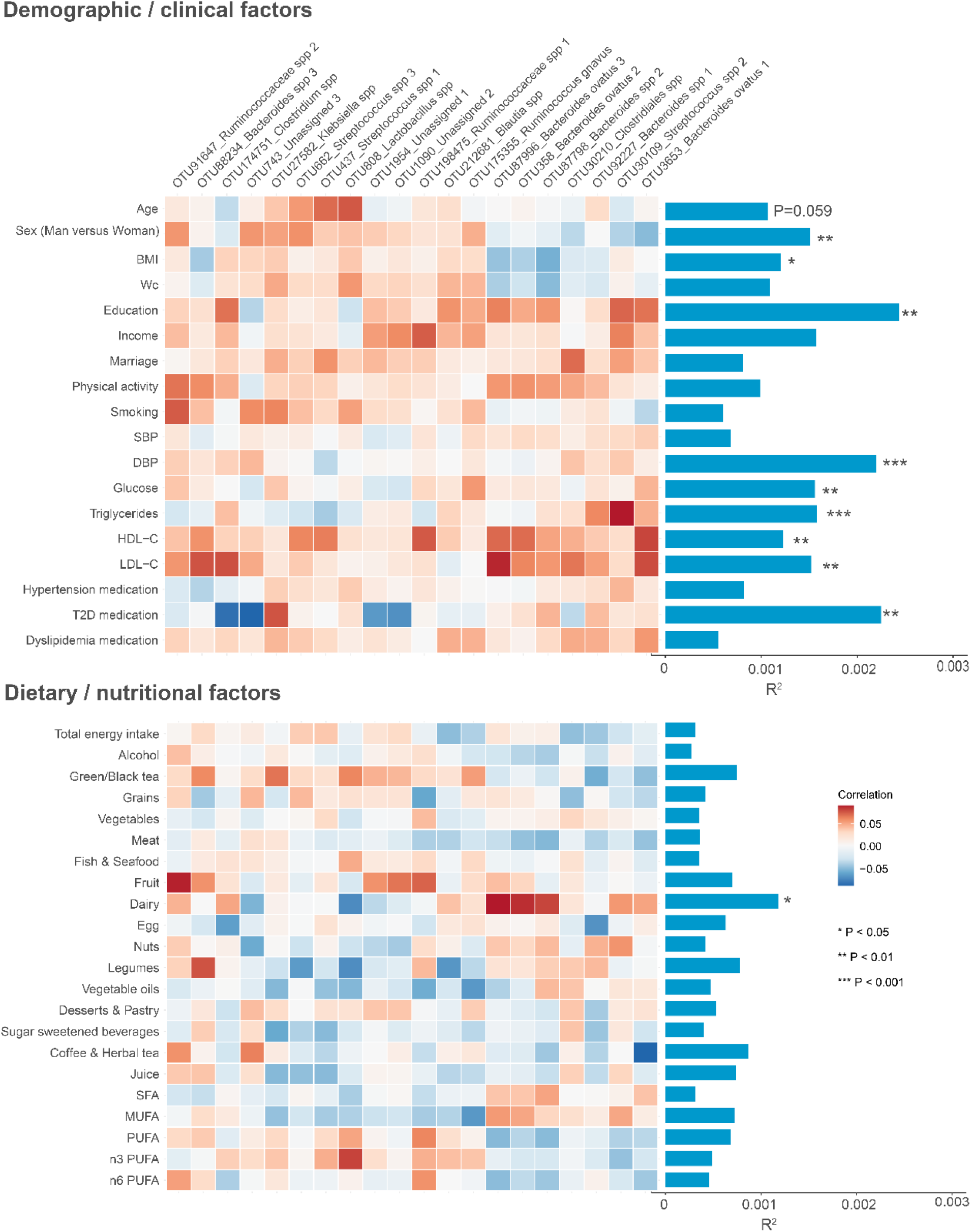
Host and environmental factors modulate the blood proteomic risk score-related core microbial OTUs. Host and environmental factors including 18 demographic/clinical items and 22 dietary/nutritional items were used in this analysis (n=1729). The bar plot indicates the explained variation of the core OTUs composition (Bray-Curtis distance) by each item. The heatmap next to the bar plot shows the correlation coefficients of each item with the core OTUs.

## Discussion

Our findings suggest that, among healthy non-infected individuals, gut microbial features are highly predictive of the blood proteomic biomarkers of severe COVID-19 disease. The disruption of the corresponding gut microbiome features may potentially predispose healthy individuals to abnormal inflammatory status, which may further account for the COVID-19 susceptibility and severity. The fecal metabolomics analysis reveals that amino acid-related pathway may provide the key link between the identified core gut microbiota, inflammation and COVID-19 susceptibility. Furthermore, modifications on host and environmental factors are likely to influence the above core gut microbiota compositions.

Accumulating evidence suggests that “cytokine storm”, an excessive production of inflammatory cytokines, may be an important mechanism leading to the severity and death of COVID-19 patients (Huang et al., 2020; Yang et al., 2020). Therefore, anticytokine therapy for the suppression of the hyperinflammatory status of the patients is a recommended strategy to treat severe COVID-19 patients (Mehta et al., 2020; Monteleone et al., 2020). Among the 20 proteomic predictors of severe COVID-19, several most upregulated proteins are activated acute phase proteins, including serum amyloid A-1 (SAA1), SAA2, SAA4, alpha-1-antichymotrypsin (SERPINA3), complement 6 (C6) and complement factor B (CFB) (Shen et al., 2020). These proteins may be activated together with proinflammatory cytokines such as IL-6 and TNF-α following the invasion of the SARS-CoV-2. Therefore, this set of proteomic biomarkers may serve as an important biomarker or therapeutic target for treating SARS-CoV-2 infection. Beyond the previous data from the COVID-19 patients, our current study based on data from healthy non-infected participants consistently supports that the proteomic biomarkers (integrated into a score) are positively associated with proinflammatory cytokines, especially among those with an older age. These results imply that the proteomic changes my precede the progression of COVID-19 to severe phase. Moreover, our finding of more significant associations between PRS and proinflammatory cytokines among older people agree with the observation during COVID-19 outbreak that older individuals are more susceptible to the virus, leading to severity of the disease, due to the induced hyperinflammation or “cytokine storm” (Chen et al., 2020; Zhou et al., 2020).

In the present study, the core gut microbial features (20 OTUs), with a satisfied performance, outperform demographic characteristics and laboratory tests in predicting the blood proteomic biomarkers, which highlights a potential role of gut microbiota in regulating the susceptibility of COVID-19 among normal individuals. In fact, maintaining gut homeostasis has been suggested as a treatment option in the “Diagnosis and Treatment Plan of Corona Virus Disease 2019 (Tentative Sixth Edition)” issued by National Health Commission of China, as to keep the equilibrium for intestinal microecology and prevent secondary bacterial infection (National Health Commission (NHC) of the PRC, 2020). Growing evidence has shown that microbiota plays a fundamental role on the induction, training and function of the host immune system, and the composition of the gut microbiota and its activity are involved in production of inflammatory cytokines (Belkaid and Hand, 2014; Cani and Jordan, 2018). Prior studies reported that *Lactobacillus* genus was positively associated with IL-6 and IFN-γ, while *Blautia* genus was positively associated with IL-10 (Jiang et al., 2012; Pohjavuori et al., 2004; Yoshida et al., 2001); these relationships were replicated in our study. Besides, we found the PRS-related OTUs belonging to *Bacteroides* genus and *Streptococcus* genus were negatively associated with most proinflammatory factors. These results further support the reliability of the selected core OTUs.

Fecal metabolomics analyses for the identified core gut microbial OTUs suggest that these OTUs may be closely associated with amino acid metabolism, especially aminoacyl-tRNA biosynthesis pathway, arginine biosynthesis pathway, and valine, leucine and isoleucine biosynthesis pathway. As metabolic stress pathways and nutrient availability instruct immunity, amino acid levels in the tissue microenvironment are central to the maintenance of immune homeostasis (Murray, 2016). Amino acid insufficiency will cause depletion of available aminoacylated tRNA, which is essential for the host to sense amino acid limitation and immune response (Brown et al., 2016, 2010; Harding et al., 2003). A recent study on several mammalian cell models reported that when aminoacyl-tRNA synthetase was inhibited, the cytokine stimulated proinflammatory response would be substantially suppressed, and a single amino acid depletion, such as arginine or histidine, could also suppress the cytokine induced immune response (Kim et al., 2020). Thus the identified pathways regulating in aminoacyl-tRNA biosynthesis and arginine biosynthesis may be both involved in the inflammatory response. Additionally, arginine and BCAAs (i.e., valine, leucine and isoleucine), were also reported regulating innate and adaptive immune responses and enhancing intestinal development (Zhang et al., 2017). Collectively, these key roles that amino acids play in the immunoregulation may help explain how the PRS-related core OTUs modulate host inflammation via amino acid metabolism. Furthermore, given the high expression of ACE2 in the ileum and colon, and the role of ACE2 as a key regulator of dietary amino acid homeostasis and innate immunity (Hashimoto et al., 2012; Zhang et al., 2020), ACE2 may be another key mediator between gut microbiota and host inflammation. However, whether and how ACE2 may mediate the association between gut microbiota and COVID-19 severity warrants further mechanistic study.

We observed that several host demographic and clinical factors had a strong effect on the identified core OTU composition, among which drug use and metabolic phenotypes had been widely reported correlating with gut microbiome composition (Cabreiro et al., 2013; Gilbert et al., 2018; Vich Vila et al., 2020). Although these observations were quite crude, it gave us an overview of the potential influence of host and environmental factors on the PRS-related gut microbiota matrix. Those known factors contributed to the COVID-19 susceptibility also contributed to the variance of the gut microbiota, including age, sex, and indicators of clinical comorbidities (blood pressure, glucose triglycerides, high-density and low-density lipoprotein cholesterol, and diabetes medication).

In summary, our study provides novel insight that gut microbiota may underlie the susceptibility of the healthy individuals to the COVID-19. In the global crisis of COVID-19, a wide disparity in the susceptibility of the disease or disease progression has been observed. Our results provide important evidence and suggestions about the potential biological mechanism behind the diverse susceptibility among different groups of people. The discovered core gut microbial features and related metabolites may serve as a potential preventive/treatment target for intervention especially among those who are susceptible to the SARS-CoV-2 infection. They could also serve as potential therapeutic targets for drug development.

## Data Availability

The raw data of 16 S rRNA gene sequences are available at CNSA (https://db.cngb.org/cnsa/) of CNGBdb at accession number CNP0000829.

## Acknowledgements

This study was funded by the National Natural Science Foundation of China (81903316, 81773416, 81972492, 21904107, 81672086), Zhejiang Ten-thousand Talents Program (101396522001), Zhejiang Provincial Natural Science Foundation for Distinguished Young Scholars (LR19C050001), the 5010 Program for Clinical Researches (2007032) of the Sun Yat-sen University, Hangzhou Agriculture and Society Advancement Program (20190101A04), and Tencent foundation (2020). The funder had no role in study design, data collection and analysis, decision to publish, or writing of the manuscript. We thank Dr. C.R. Palmer for his invaluable comments to this study; and Westlake University Supercomputer Center for assistance in data storage and computation; and all the patients who consented to donate their clinical information and samples for analysis; and all the medical staff members who are on the front line fighting against COVID-19.

## Author Contributions

Conceptualization, J.S.Z.; Methodology, W.G. and Y.F.; Formal Analysis, W.G., Y.F., L.Y., G.D.C, X.C., M.S., and F.X.; Investigation, M.L.X., B.S, X.W., H.Z., and W.H.L.; Data curation, X.Y., H.C., Y.Z., Z.J., Z.M. and C.X.; Resources, Y.M.C., T.G., J.S.Z; Writing, Y.F. and J.S.Z.; Writing-Review & Editing, J.S.Z, T.G., J.W, Y.F. and W.L.G; Visualization, M.S. and F.X.; Supervision, J.S.Z., Y.M.C., and T.G.; Funding Acquisition, J.S.Z., Y.M.C. and T.G.

## STAR MEHTODS

### RESOURCE AVAILABILITY

#### Lead Contact

Further information and requests for resources and reagents should be directed to and will be fulfilled by the Lead Contact, Ju-Sheng Zheng.

#### Materials Availability

This study did not generate new unique reagents.

### SUBJECT DETAILS

#### COVID-19 proteomics data set

Detailed information about the COVID-19 patients and proteomics data set is described in our recent publication (Shen et al., 2020). Briefly, the proteome of sera from 46 COVID-19 patients and 53 control samples from Taizhou Public Health Medical Center were analyzed by TMTpro 16 plex-based quantitative proteomics technology. All the patients were diagnosed between January 23 and February 4, 2020. According to the Chinese Government Diagnosis and Treatment Guideline for COVID-19, the COVID-19 patients were classified into four groups, (1) mild (mild symptoms without pneumonia); (2) typical (fever or respiratory tract symptoms with pneumonia); (3) severe (fulfill any of the three criteria: respiratory distress, respiratory rate≥30 times/min; mean oxygen saturation ≤ 93% in resting state; arterial blood oxygen partial pressure/oxygen concentration ≤ 300mmHg); and (4) critical (fulfill any of the three criteria: respiratory failure and require mechanical ventilation; shock incidence; admission to ICU with other organ failure). We treated mild and typical patients as a non-severe COVID-19 group, and the others a severe COVID-19 group.

#### Healthy subjects, sample collection, and clinical metadata

In the present study, the healthy (non-infected) subjects are from the community-based Guangzhou Nutrition and Health Study (GNHS), and the detailed study designs of GNHS have been reported previously (Zhang et al., 2014). Briefly, participants were enrolled between 2008 and 2013, and followed up to May 2018. Blood samples were collected at enrollment and follow-up visits, and stool samples were collected only during follow-up visits. All the blood samples were collected as venous whole blood in the early morning before diet using serum separation tubes. The blood samples were centrifuged at 3,500 rpm for 10 min for serum collection. The serum samples were frozen at -80°C. The stool samples were collected at a local study site within the School of Public Health at Sun Yat-sen University, and were transferred to a -80°C facility within 4 hours after collection.

Demographic and lifestyle factors were all collected by questionnaire during on-site face-to-face interviews. Habitual dietary intakes over the past 12 months were assessed by a food frequency questionnaire, as previously described (Zhang CX, 2009). Physical activity was assessed as a total metabolic equivalent for task (MET) hours per day on the basis of a validated questionnaire for physical activity (Liu et al., 2001).

Anthropometric factors were measured by trained nurses on site during the baseline interview. Fasting venous blood samples were taken at each recruitment or follow-up visit. Serum low-density lipoprotein cholesterol and glucose were measured by coloimetric methods using a Roche Cobas 8000 c702 automated analyzer (Roche Diagnostics GmbH, Shanghai, China). Intra-assay coefficients of variation (CV) was 2.5% for glucose. Insulin was measured by electrochemiluminescence immunoassay (ECLIA) methods using a Roche cobas 8000 e602 automated analyzer (Roche Diagnostics GmbH, Shanghai, China). High-performance liquid chromatography was used to measure glycated hemoglobin (HbA1c) using the Bole D-10 Hemoglobin A1c Program on a Bole D-10 Hemoglobin Testing System, and the intraassay CV was 0.75%.

#### Ethics

This study has been approved by the Ethical/Institutional Review Board of Taizhou Public Health Medical Center, the Ethics Committee of the School of Public Health at Sun Yat-sen University and Ethics Committee of Westlake University.

### METHOD DETAILS

#### Proteomic analysis

1 μL of serum sample from each patient was analyzed using proteomics technology. The serum was firstly denatured with 20 µL of buffer containing 8 M urea (Sigma, #U1230) in 100 mM ammonium bicarbonate at 30°C for 30 min. The lysates were reduced with 10 mM tris (2-carboxyethyl) phosphine (TCEP, Sigma #T4708) at room temperature for 30 min, and were then alkylated with 40 mM iodoacetamide (IAA, Sigma, #SLCD4031) in darkness for 45 min. The solution was then diluted with 70 µL 100 mM ammonium bicarbonate to make sure urea concentration is less than 1.6M, and was subjected to two times of tryptic digestion (Hualishi Tech. Ltd, Beijing, China), each step with 2.5 μL trypsin (0.4 μg/μL), at 32°C for 4 hr and 12 hr, respectively. Thereafter, the solution was acidified with 1% trifluoroacetic (TFA) (Thermo Fisher Scientific, #T/3258/PB05) to pH 2–3 to stop the reaction. Peptides were cleaned using C18 (Thermo, #60209-001).

Peptide samples were then injected for LC-MS/MS analysis using an Eksigent NanoLC 400 System (Eksigent, Dublin, CA, USA) coupled to a TripleTOF 5600 system (SCIEX, CA, USA). Briefly, peptides were loaded onto a trap column (5 µm, 120 Å, 10 × 0.3 mm), and were separated along a 20 min LC gradient (5–32% buffer B, 98% ACN, 0.1% formic acid in HPLC water; buffer A: 2% ACN, 0.1% formic acid in HPLC water) on an analytical column (3 µm, 120 Å, 150 × 0.3 mm) at a flow rate of 5 µL/min. The SWATH-MS method is composed of a 100 ms of full TOF MS scan with the acquisition range of 350-1250 *m/z*, followed by MS/MS scans performed on all precursors (from 100 to 1500 Da) in a cyclic manner (Gillet et al., 2012). A 55-variable-Q1 isolation window scheme was used in this study. The accumulation time was set at 30 ms per isolation window, resulting in a total cycle time of 1.9 s.

After SWATH acquisition, the Wiff files were converted into mzXML format using msconvert (ProteoWizard 3.0) (Kessner et al., 2008) and analyzed using OpenSWATH (2.1) (Kessner et al., 2008) against a pan human spectral library (Kessner et al., 2008) that contains 43899 peptide precursors and 1667 unique Swiss-Prot proteins protein groups. The retention time extraction window was set at 120 seconds, and the *m/z* extraction was performed with 30 ppm tolerance. Retention time was then calibrated using Common internal Retention Time standards (CiRT) peptides (Kessner et al., 2008). Peptide precursors were identified by OpenSWATH (version 2.0) and pyprophet (version 0.24) with FDR<0.01 to quantify the proteins in each sample.

#### Measurement of inflammatory biomarkers

For samples collected at baseline, Human FlowCytomix (Simplex BMS8213FF and BMS8288FF, eBioscience, San Diego, CA, USA) and the Human Basic Kit FlowCytomix (BMS8420FF, eBioscience, San Diego, CA, USA) on a BD FACSCalibur instrument (BD Biosciences, Franklin Lakes, NJ, USA) were used for the measurements of serum tumor necrosis factor (TNF-α), Interleukin-6 (IL-6), and Interleukin-1β (IL-1β). High-sensitivity CRP was measured using a [Cardiac C-Reactive Protein (Latex) High Sensitive (CRPHS) kit], and detected on a Cobas c701 automatic analyzer. The between-plate CVs were 14.1% for MCP1, 6.6% for TNF-α, 2.5% for IL-6, and 10.2% for IL-1β.

For samples collected during follow-up visits, serum cytokine levels were assessed by electrochemiluminescence based immunoassays using the MSD V-Plex Proinflammatory Panel 1 (human) kit. 50 µL of serum derived from whole blood by centrifugation (10 min,3,500 rpm) was processed according to the manufacturer’s instructions. Briefly, the serum was diluted at a minimum of 2-fold dilution at first, while the detection antibodies were combined and added to 2400 µL of diluent. Thereafter, wash buffer and read buffer T were prepared as instructed. After finishing washing and adding samples, washing and adding detection antibody solution and washing plates again, then the plate could be analyzed on an MSD instrument. Accuracy and precision are evaluated by measuring calibrators across multiple runs and multiple lots. Intra-run coefficient of variations (CVs) are typically below 7% and inter-ran CVs are typically below 15%. In the present study, the inters-run CVs of calibrators were 2.2% for IL-1β, 2.5% for IL-2, 1.6% for IL-4, 1.7 for IL-6, 3.6 for IL-8, 2.5 for 1.3% for IL-10, 1.1% for IL-12p70, 0.87% for IL-13, TNF-α, 2.0% for IFN-γ.

#### Microbiome analysis ---- DNA extraction

Total bacterial DNA was extracted using the QIAamp® DNA Stool Mini Kit (Qiagen, Hilden, Germany) following the manufacturer’s instructions. DNA concentrations were measured using the Qubit quantification system (Thermo Scientific, Wilmington, DE, US). The extracted DNA was then stored at -20 °C.

#### Microbiome analysis ---- 16S rRNA gene amplicon sequencing

The 16S rRNA gene amplification procedure was divided into two PCR steps, in the first PCR reaction, the V3-V4 hypervariable region of the 16S rRNA gene was amplified from genomic DNA using primers 341F(CCTACGGGNGGCWGCAG) and 805R(GACTACHVGGGTATCTAATCC). Amplification was performed in 96-well microtiter plates with a reaction mixture consisting of 1X KAPA HiFi Hot start Ready Mix, 0.1µM primer 341 F, 0.1 µM primer 805 R, and 12.5 ng template DNA giving a total volume of 50 µL per sample. Reactions were run in a T100 PCR thermocycle (BIO-RAD) according to the following cycling program: 3 min of denaturation at 94 °C, followed by 18 cycles of 30 s at 94 °C (denaturing), 30 s at 55 °C (annealing), and 30 s at 72 °C (elongation), with a final extension at 72 °C for 5 min. Subsequently, the amplified products were checked by 2% agarose gel electrophoresis and ethidium bromide staining. Amplicons were quantified using the Qubit quantification system (Thermo Scientific, Wilmington, DE, US) following the manufacturers’ instructions. Sequencing primers and adaptors were added to the amplicon products in the second PCR step as follows 2 µL of the diluted amplicons were mixed with a reaction solution consisting of 1×KAPA HiFi Hotstart ReadyMix, 0.5µM fusion forward and 0.5µM fusion reverse primer, 30 ng Meta-gDNA(total volume 50 µL). The PCR was run according to the cycling program above except with cycling number of 12. The amplification products were purified with Agencourt AMPure XP Beads (Beckman Coulter Genomics, MA, USA) according to the manufacturer’s instructions and quantified as described above. Equimolar amounts of the amplification products were pooled together in a single tube. The concentration of the pooled libraries was determined by the Qubit quantification system. Amplicon sequencing was performed on the Illumina MiSeq System (Illumina Inc., CA, USA). The MiSeq Reagent Kits v2 (Illumina Inc.) was used. Automated cluster generation and 2 × 250 bp paired-end sequencing with dual-index reads were performed.

#### Microbiome analysis ---- 16S rRNA gene sequence data processing

Fastq-files were demultiplexed by the MiSeq Controller Software (Illumina Inc.). The sequence was trimmed for amplification primers, diversity spacers, and sequencing adapters, merge-paired and quality filtered by USEARCH. UPARSE was used for OTU clustering equaling or above 97%. Taxonomy of the OTUs was assigned and sequences were aligned with RDP classifier. The OTUs were analyzed by phylogenetic and operational taxonomic unit (OTU) methods in the Quantitative Insights into Microbial Ecology (QIIME) software version 1.9.0 (Caporaso et al., 2010).

#### Metabolomic analysis ---- sample preparation and instrumentation

Targeted metabolomics approach was used to analyze fecal samples, with a total of 198 metabolites quantified. Feces samples were thawed on ice-bath to diminish degradation. About 10mg of each sample was weighed and transferred to a new 1.5mL tube. Then 25μL of water was added and the sample was homogenated with zirconium oxide beads for 3 minutes. 185μL of ACN/Methanol (8/2) was added to extract the metabolites. The sample was centrifuged at 18000g for 20 minutes. Then the supernatant was transferred to a 96-well plate. The following procedures were performed on a Biomek 4000 workstation (Biomek 4000, Beckman Coulter, Inc., Brea, California, USA). 20μL of freshly prepared derivative reagents was added to each well. The plate was sealed and the derivatization was carried out at 30°C for 60 min. After derivatization, 350μL of ice-cold 50% methanol solution was added to dilute the sample. Then the plate was stored at -20°C for 20 minutes and followed by 4000g centrifugation at 4 °C for 30 minutes. 135μL of supernatant was transferred to a new 96-well plate with 15μL internal standards in each well. Serial dilutions of derivatized stock standards were added to the left wells. Finally the plate was sealed for LC-MS analysis.

An ultra-performance liquid chromatography coupled to tandem mass spectrometry (UPLC-MS/MS) system (ACQUITY UPLC-Xevo TQ-S, Waters Corp., Milford, MA, USA) was used to quantitate the microbial metabolite in the present study. The optimized instrument settings are briefly described below. ACQUITY UPLC BEH C18 1.7 µM VanGuard pre-column (2.1×5 mm) and ACQUITY UPLC BEH C18 1.7 µM analytical column (2.1 × 100 mm) were used. The column temperature was 40°C and sample manager temperature was 10°C. Mobile phase A was water with 0.1% formic acid, and B was acetonitrile / IPA (90:10). The gradient conditions were as follows: 0-1 min (5% B), 1-12 min (5-80% B), 12-15 min (80-95% B), 15-16 min (95-100%B), 16-18 min (100%B), 18-18.1 min (100-5% B), 18.1-20 min (5% B), at a flow rate of 0.40 mL/min. The capillary of mass spectrometer were 1.5 (ESI+) and 2.0 (ESI-), while the source temperature and desolvation temperature was 150°C and 550°C, respectively. The desolvation gas flow was 1000 L/hour.

#### Metabolome analysis ---- Analytical quality control procedures

The rapid turnover of many intracellular metabolites makes immediate metabolism quenching necessary. The extraction solvents are stored in -20°C freezer overnight and added to the samples immediately after the samples were thawed. We use ice-salt bath to keep the samples at a low temperature and minimize sample degradation during sample preparation. All the prepared samples should be analyzed within 48 hours after sample extraction and derivatization.

A comprehensive set of rigorous quality control/assurance procedures is employed to ensure a consistently high quality of analytical results, throughout controlling every single step from sample receipt at laboratory to final deliverables. The ultimate goal of QA/QC is to provide the reliable data for biomarker discovery study and/ or to aid molecular biology research. To achieve this, three types of quality control samples i.e., test mixtures, internal standards, and pooled biological samples are routinely used in the metabolomics platform. In addition to the quality controls, conditioning samples, and solvent blank samples are also required for obtaining optimal instrument performance.

Test mixtures comprise a group of commercially available standards with a mass range across the system mass range used for the study samples. These samples were analyzed at the beginning and end of each batch run to ensure that the instruments were performing within laboratory specifications (retention time stability, chromatographic peak shape, and peak signal intensity). The retention time shift should be within 4 sec. and the difference of peak intensity should be within 15% for LC-MS.

Internal standards were added to the test samples in order to monitor analytical variations during the entire sample preparation and analysis processes. The Pooled QC samples were prepared by mixing aliquots of the study samples such that the pooled samples broadly represent the biological average of the whole sample set. The QC samples for this project were prepared with the test samples and injected at regular intervals (after every 14 test samples for LC-MS) throughout the analytical run.

Reagent blank samples are a mixture of solvents used for sample preparation and are commonly processed using the same procedures as the samples to be analyzed. The reagent blanks serve as a useful alert to systematic contamination. As the reagent blanks consist of high purity solvents and are analyzed using the same methods as the study samples, they are also used to wash the column and remove cumulative matrix effects throughout the study.

The calibrators consist of a blank sample (matrix sample processed without internal standard), a zero sample (matrix sample processed with internal standard), and a series of seven concentrations covering the expected range for the metabolites present in the specific biological samples. LLOQ and ULOQ are the lowest and highest concentration of the standard curve that can be measured with acceptable accuracy and precision.

To diminish analytical bias within the entire analytical process, the samples were analyzed in group pairs but the groups were analyzed randomly. The QC samples, calibrators, and blank samples were analyzed across the entire sample set.

#### Metabolome analysis ---- Software and quantitation

The raw data files generated by UPLC-MS/MS were processed using the QuanMET software (v2.0, Metabo-Profile, Shanghai, China) to perform peak integration, calibration, and quantification for each metabolite. The current QuanMET is hosted on Dell PowerEdge R730 Servers operated with Linux Ubuntu 16.10 OS. The secured Java UI (User Interface) permits the user have access to use a great variety of statistical tools for viewing and exploring project data.

Mass spectrometry-based quantitative metabolomics refers to the determination of the concentration of a substance in an unknown sample by comparing the unknown to a set of standard samples of known concentration (i.e., calibration curve). The calibration curve is a plot of how the analytical signal changes with the concentration of the analyte (the substance to be measured). For most analyses a plot of instrument response vs. concentration will show a linear relationship and the concentration of the measured samples were calculated.

## STATISTICAL ANALYSIS

### Dataset at each step of analyses among the healthy individuals from Guangzhou Nutrition and Health Study

#### Dataset 1 (n=990)

data from the baseline of GNHS. A total of 990 subjects with measurement of serum proteomics at baseline of the GNHS cohort were included in the initial discovery cohort. Among this initial discovery cohort, 455 subjects had data of serum IL-1β and IL-6, 456 subjects had data of serum TNF-α, and 953 subjects had serum hsCRP data. These data were used to investigate the relationship between the PRS and host inflammatory status.

#### Dataset 2 (n=301)

data from a follow-up visit of GNHS. A sub-cohort of 301 participants with measurement of both gut microbiota (16s rRNA) and blood proteomics data. Gut microbiota data were collected and measured during a follow-up visit of the cohort participants, with a cross-sectional subset of the individuals (n=132) having blood proteomic data at the same time point as the stool collection and another independent prospective subset of the individuals (n=169) having proteomic data at a next follow-up visit ∼3 years later than the stool collection. Data from these subjects were used to explore the predictive capacity of the gut microbiota for PRS.

#### Dataset 3 (n=366)

data from a follow-up visit of GNHS. Independent from the above 301 subjects in dataset 2, there were additionally 336 subjects with both fecal 16s rRNA sequencing and serum inflammatory cytokines data, at the same time point during follow-up. The data from these 336 subjects were used to examine the relationships between the core OTUs and 10 host inflammatory cytokines.

#### Dataset 4 (n=987)

data from a follow-up visit of GNHS. A total of 987 individuals had received fecal metabolomics and fecal microbiome examination at the same time point during the follow-up visit. These subjects were included in the analysis to assess the relationships between the core gut microbiota and fecal metabolomics.

#### Dataset 5 (n=1729)

data from a follow-up visit of GNHS. In total, 1729 participants finished food frequency questionnaire, demographic questionnaire and medical examination, and provided stool samples during follow-up. Thus, this subset of 1729 subjects were included to test how the dietary habits, lifestyle and health status influence the gut microbiota composition.

In summary, a sum of 2413 healthy non-infected individuals are involved in the present study, which mainly consists of a subset of subjects with proteomic data at baseline (n=990) and a subset of subjects with gut microbiome and metabolome data at a follow-up visit (n=2172, within which 301 individuals also had proteomic data).

### Data imputation and presentation

Missing values in proteomic features were imputed with 50% of the minimal value. Data are presented as mean ± SD or percentage as indicated. Statistical tests used to compare conditions are indicated in figure legends. Unless otherwise stated, statistical analysis was performed using Python 3.7, R software (version 3.6.1, R foundation for Statistical Computing, Austria), and Stata 15 (StataCorp, College Station, TX, USA).

### Construction of proteomic risk score (*PRS*)

We used 20 out of 22 previously identified proteomic biomarkers to construct a proteomic risk score (PRS) for severe COVID-19 in COVID-19 patients and healthy participants.

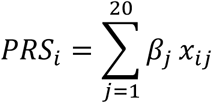

Where, *PRS*_*i*_ is a proteomic risk score for individual *i*, 20 is the number of proteins involved the score construction, *x*_*ij*_ is the Z score of abundance of the protein *j* for individual *i. β* is 1 or -1 depending on the association between the protein *j* and risk of progressing to clinically severe phase (1, up-regulated in severe patients, -1, down-regulated in severe patients).

### Association of PRS with the risk of progressing to clinically severe phase

Poisson regression model was used to examine the association of PRS with the risk of progressing to clinically severe phase among 31 COVID-19 patients (18 non-severe patients; 13 severe patients), adjusting for age, sex and BMI.

### Correlation between PRS and pro-inflammatory biomarkers

Spearman correlation analysis was used to examine the correlation between PRS and pro-inflammatory biomarkers (i.e., hsCRP, IL-1β, IL-6 and TNF-α). p<0.05 was considered as statistically significant.

### Machine learning algorithms for identifying microbial features to predict PRS

A 10-fold cross-validation (CV) implementation of gradient boosting framework —LightGBM and SHAP (Shapley Additive exPlanations) was used to link input gut microbial features with PRS (Ke et al., 2017; Lee, 2017). A 10-fold CV predict implementation was used to generate a OTU-predicted PRS value for each participant. In this approach, each LightGBM model is trained on 90% of the cohort with 10-fold CV, and PRS is predicted for the 10% of the participants who were not used for model optimization. This process is repeated ten-fold resulting in a test PRS set for each participant and ten different average absolute SHAP value for each OTUs. The top 20 ranked OTUs based the sum of the average absolute SHAP value across ten-fold were included in further analysis. The R^2^ score was computed by taking the mean of all the R^2^ scores across the 10 out-of-sample predictions. Pearson r was calculated using actual PRS and predicted PRS for the entire cohort. We also compared the predictive performance for the top 20 ranked OTUs, demographic characteristics and laboratory tests (age, BMI, sex, blood pressure and blood lipids). Our predictor is based on code adapted from the sklearn 0.15.2 lightgbm regression (Pedregosa et al., 2011).

### The relationship between the identified core OTUs and host inflammatory cytokines

Spearman correlation analysis was used to examine the correlation between PRS and cytokines (i.e., IL-1β, IL-2, IL-4, IL-6, IL-8, IL-10, IL-12p70, IL-13, TNF-α And IFN-γ). p<0.05 was considered as statistically significant.

### Relationship between OTUs and fecal metabolites

Prior to the analysis, we excluded the participants with T2D medication use, and all fecal metabolites were natural logarithmic transformed to reduce skewness of traits distributions. Similarly, to reduce skewness of the distribution of microbial taxa counts, we first added 1 to all OTUs and then performed natural log transformation. The relationship between fecal metabolites and microbial OTUs was assessed by linear regression analysis while adjusting for age, sex, BMI. Multiple testing was adjusted using Benjamini and Hochberg method, with a false discovery rate (FDR) of <0.05 being considered statistically significant. Metabolites showed significant associations with more than half of the selected microbial OTUs, were used for subsequent pathway analysis using MetaboAnalyst 4.0 (Chong et al., 2019).

### Associations of host and environmental factors with gut microbial features

We assessed how many variations in the identified core OTUs composition (Bray-Curtis distance) can be explained by host and environmental factors (40 factors) using the function *adonis* from the R package vegan. The p value was determined by 1000x permutations. The total variation explained was also calculated per category (demographic/clinical category and dietary/nutritional factors) and for all factors together. Spearman correlation analysis was used to assess the potential effect of each factor on each of the core OTU. Multiple testing was adjusted using Benjamini and Hochberg method, with a false discovery rate (FDR) of <0.05 being considered statistically significant.

## Supplementary Figure Legends

**Figure S1.**
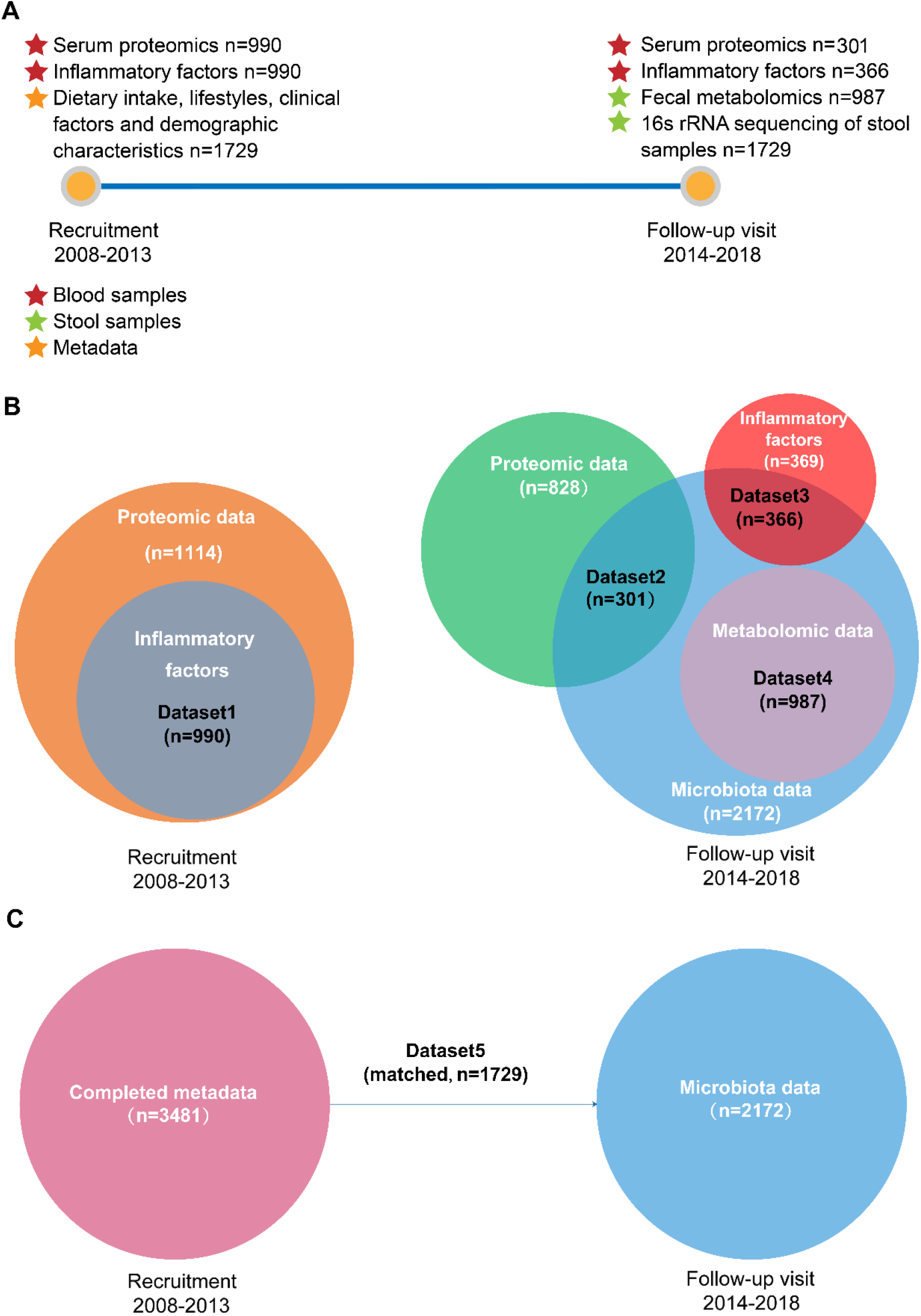
Timeline of the participants enrollment, follow-up visit and sample collection in the Guangzhou Health and Nutrition Study. **(A)** At baseline, 4048 subjects provided completed metadata (required for analysis in the present study), and 1114 subjects provided blood samples (1114 for proteomic analysis and 990 for measurement of inflammatory factors). At follow-up visit, 2172 subjects provided stool samples (n=1729 for 16s rRNA sequencing; n=987 for metabolomic analysis), among which 667 subjects provided blood samples (n=301 for proteomic analysis; n=366 for measurement of inflammatory factors). (B) Detail information about the number of participants in the dataset1-dataset4 used in the present study. (C) Detail information about the number of participants in the dataset5 used in the present study.

**Figure S2.**
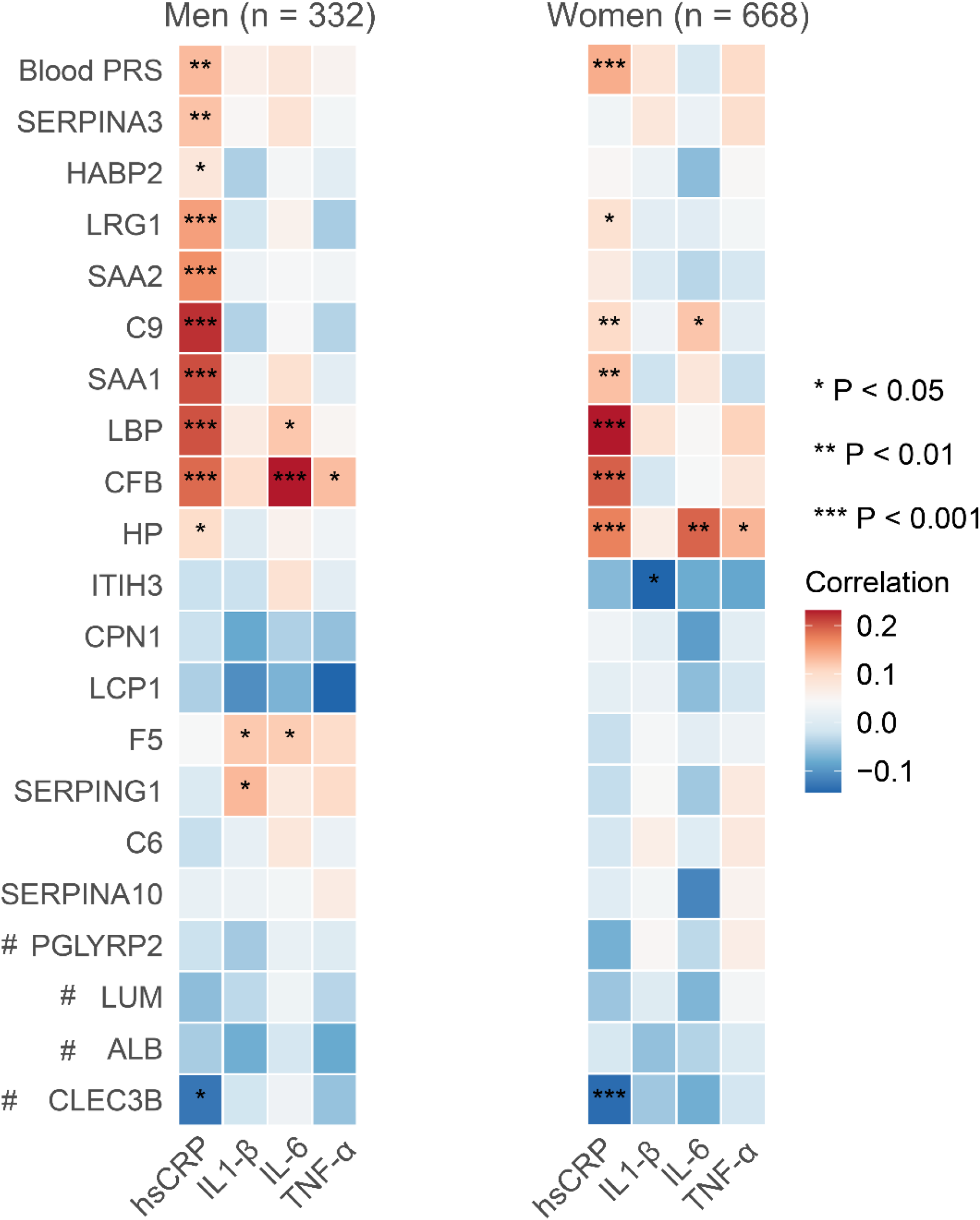
The correlation of the blood proteomic biomarkers and PRS with host inflammatory markers stratified by sex (n=990). The color of the heatmap indicates the Spearman correlation coefficients (blue-negative, red-positive). # protein down-regulated in severe patients, else, up-regulated.

**Figure S3.**
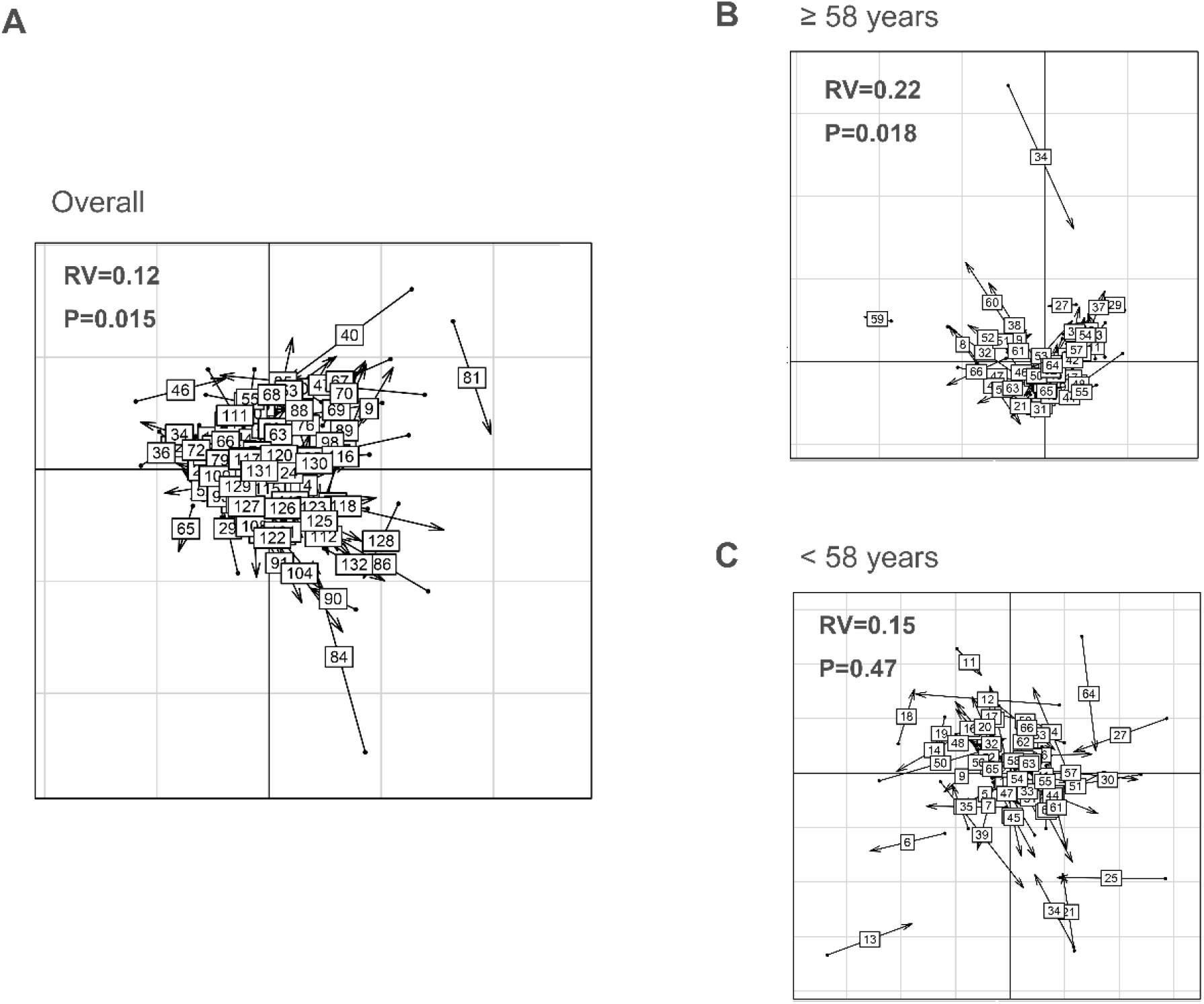
Co-variation of the core OTUs abundance and predictive proteomic biomarkers of COVID-19 (n=132). Co-inertia analysis of the relationship between 20 identified OTUs and 20 predictive proteomic biomarkers among a cross-sectional subset of 132 individuals. Each sample is represented with an arrow. The sample projection in the OTUs and the proteomic biomarkers space are represented by the starting point and the end of the arrow, respectively. Length of the arrow represents distance between the projections. (A, overall; B, age ≥ 58; C, age <58).

## Supplementary Tables

**Supplementary Table 1.**
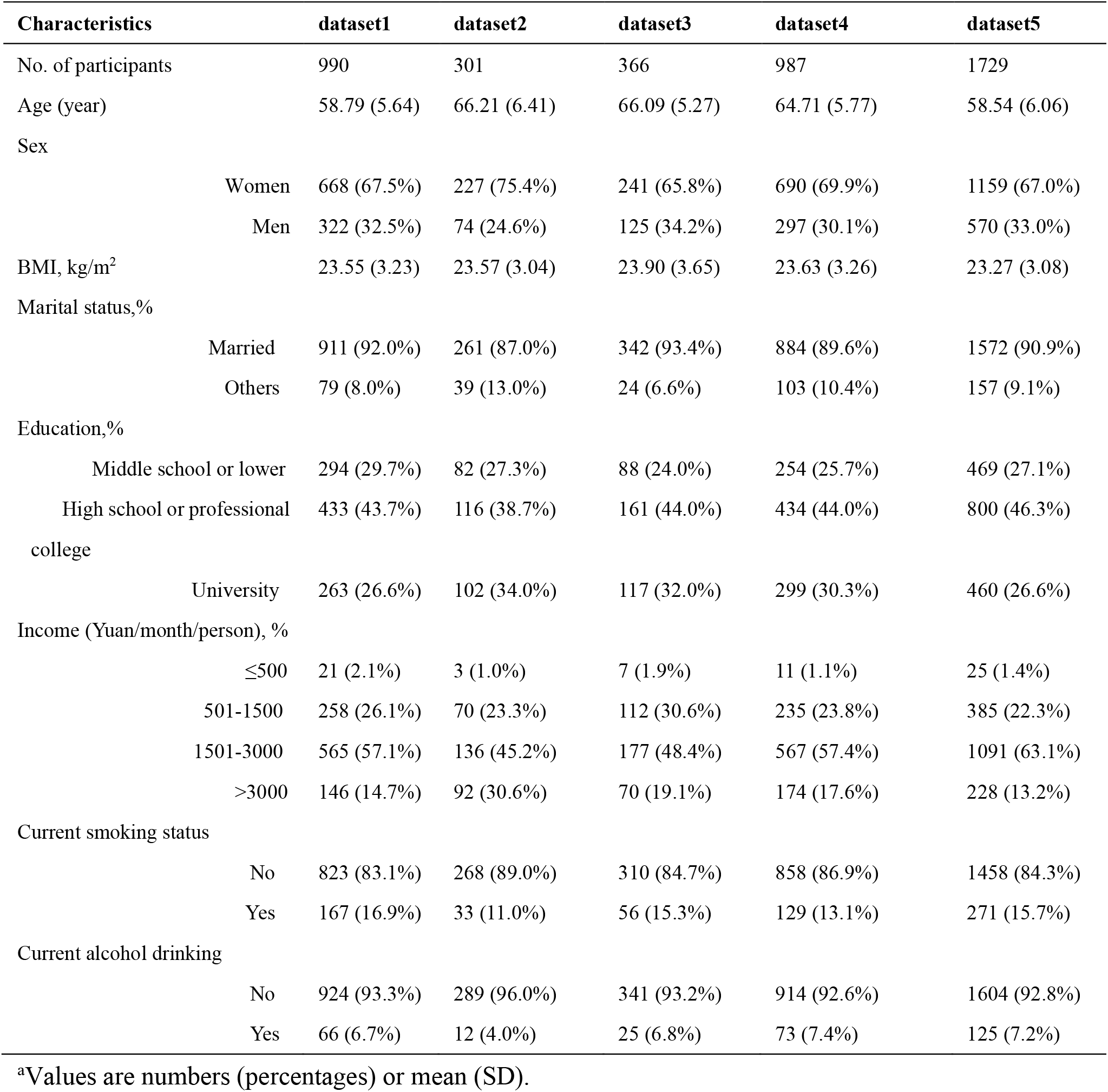
Characteristics of the data analysis set^a^

| Characteristics |  | dataset1 | dataset2 | dataset3 | dataset4 | dataset5 |
| --- | --- | --- | --- | --- | --- | --- |
| No. of participants |  | 990 | 301 | 366 | 987 | 1729 |
| Age (year) |  | 58.79 (5.64) | 66.21 (6.41) | 66.09 (5.27) | 64.71 (5.77) | 58.54 (6.06) |
| Sex |  |  |  |  |  |  |
|  | Women | 668 (67.5%) | 227 (75.4%) | 241 (65.8%) | 690 (69.9%) | 1159 (67.0%) |
|  | Men | 322 (32.5%) | 74 (24.6%) | 125 (34.2%) | 297 (30.1%) | 570 (33.0%) |
| BMI, kg/m<sup>2</sup> |  | 23.55 (3.23) | 23.57 (3.04) | 23.90 (3.65) | 23.63 (3.26) | 23.27 (3.08) |
| Marital status,% |  |  |  |  |  |  |
|  | Married | 911 (92.0%) | 261 (87.0%) | 342 (93.4%) | 884 (89.6%) | 1572 (90.9%) |
|  | Others | 79 (8.0%) | 39 (13.0%) | 24 (6.6%) | 103 (10.4%) | 157 (9.1%) |
| Education,% |  |  |  |  |  |  |
|  | Middle school or lower | 294 (29.7%) | 82 (27.3%) | 88 (24.0%) | 254 (25.7%) | 469 (27.1%) |
|  | High school or professional college | 433 (43.7%) | 116 (38.7%) | 161 (44.0%) | 434 (44.0%) | 800 (46.3%) |
|  | University | 263 (26.6%) | 102 (34.0%) | 117 (32.0%) | 299 (30.3%) | 460 (26.6%) |
| Income (Yuan/month/person), % |  |  |  |  |  |  |
|  | ≤500 | 21 (2.1%) | 3 (1.0%) | 7 (1.9%) | 11 (1.1%) | 25 (1.4%) |
|  | 501-1500 | 258 (26.1%) | 70 (23.3%) | 112 (30.6%) | 235 (23.8%) | 385 (22.3%) |
|  | 1501-3000 | 565 (57.1%) | 136 (45.2%) | 177 (48.4%) | 567 (57.4%) | 1091 (63.1%) |
|  | >3000 | 146 (14.7%) | 92 (30.6%) | 70 (19.1%) | 174 (17.6%) | 228 (13.2%) |
| Current smoking status |  |  |  |  |  |  |
|  | No | 823 (83.1%) | 268 (89.0%) | 310 (84.7%) | 858 (86.9%) | 1458 (84.3%) |
|  | Yes | 167 (16.9%) | 33 (11.0%) | 56 (15.3%) | 129 (13.1%) | 271 (15.7%) |
| Current alcohol drinking |  |  |  |  |  |  |
|  | No | 924 (93.3%) | 289 (96.0%) | 341 (93.2%) | 914 (92.6%) | 1604 (92.8%) |
|  | Yes | 66 (6.7%) | 12 (4.0%) | 25 (6.8%) | 73 (7.4%) | 125 (7.2%) |

<sup>a</sup>Values are numbers (percentages) or mean (SD).

**Supplementary Table 2:**
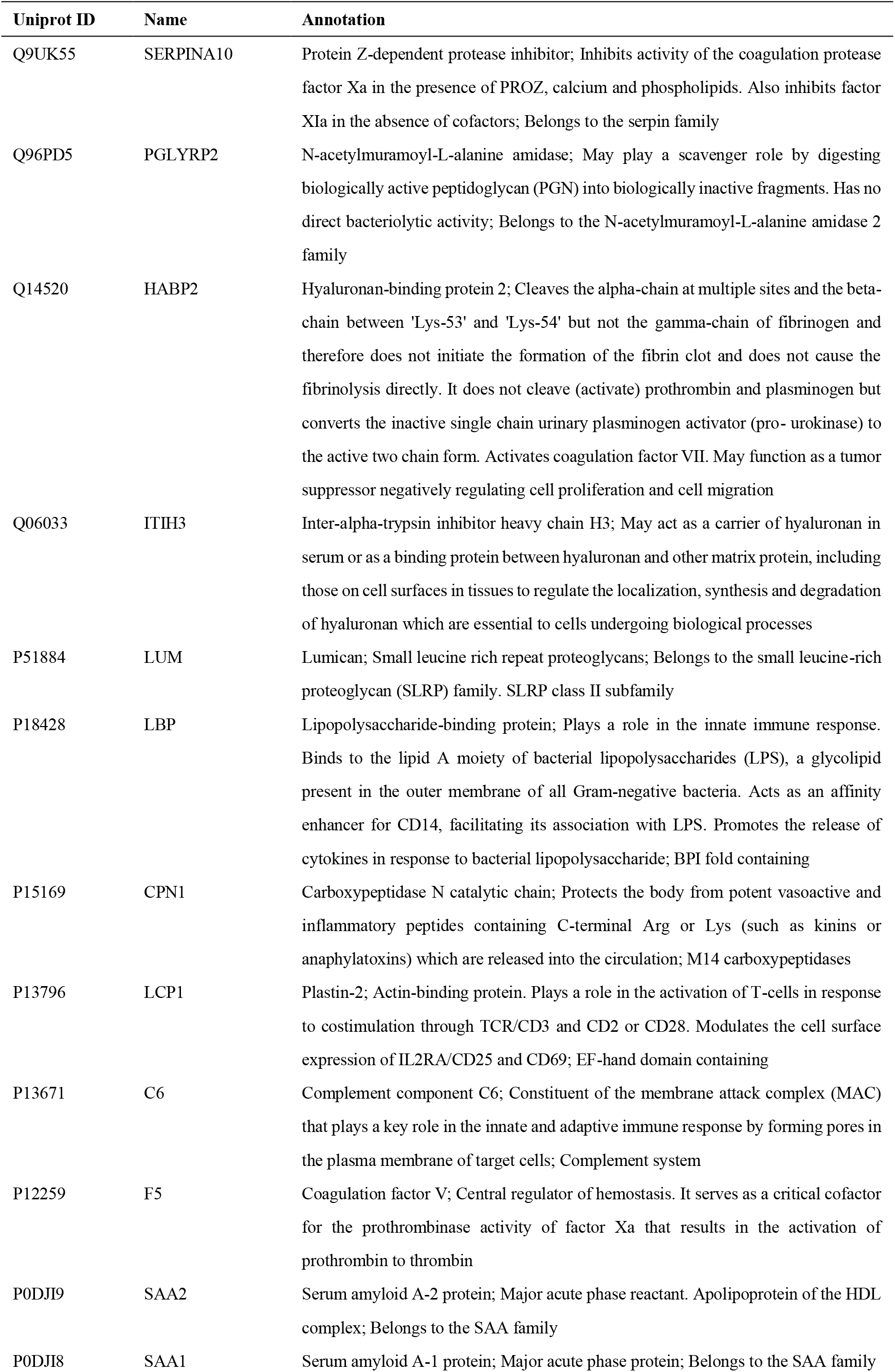
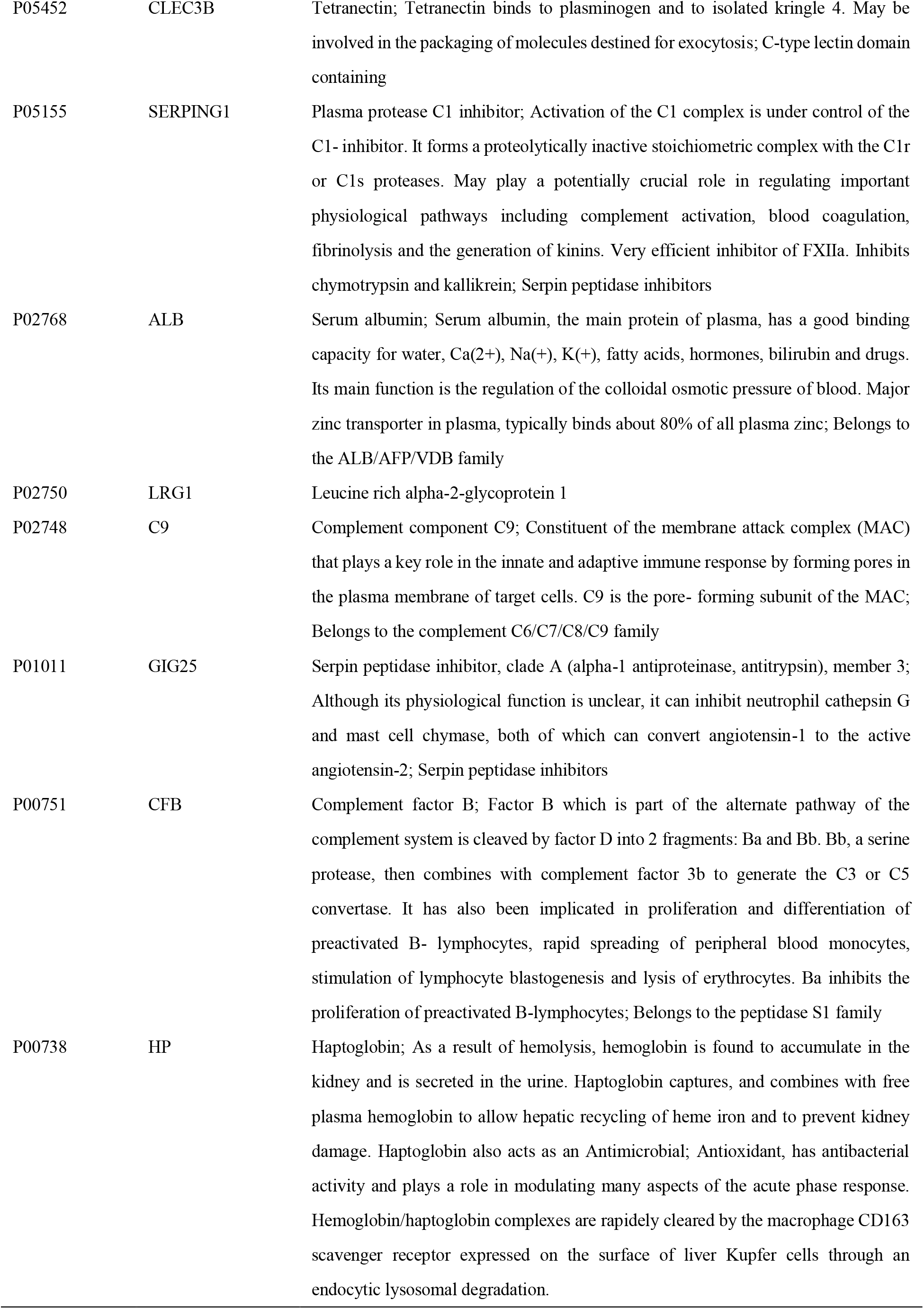
List of the 20 proteomic biomarkers integrated in the proteomic risk score

| Uniprot ID | Name | Annotation |
| --- | --- | --- |
| Q9UK55 | SERPINA10 | Protein Z-dependent protease inhibitor; Inhibits activity of the coagulation protease factor Xa in the presence of PROZ, calcium and phospholipids. Also inhibits factor XIa in the absence of cofactors; Belongs to the serpin family |
| Q96PD5 | PGLYRP2 | N-acetylmuramoyl-L-alanine amidase; May play a scavenger role by digesting biologically active peptidoglycan (PGN) into biologically inactive fragments. Has no direct bacteriolytic activity; Belongs to the N-acetylmuramoyl-L-alanine amidase 2 family |
| Q14520 | HABP2 | Hyaluronan-binding protein 2; Cleaves the alpha-chain at multiple sites and the beta-chain between 'Lys-53' and 'Lys-54' but not the gamma-chain of fibrinogen and therefore does not initiate the formation of the fibrin clot and does not cause the fibrinolysis directly. It does not cleave (activate) prothrombin and plasminogen but converts the inactive single chain urinary plasminogen activator (pro- urokinase) to the active two chain form. Activates coagulation factor VII. May function as a tumor suppressor negatively regulating cell proliferation and cell migration |
| Q06033 | ITIH3 | Inter-alpha-trypsin inhibitor heavy chain H3; May act as a carrier of hyaluronan in serum or as a binding protein between hyaluronan and other matrix protein, including those on cell surfaces in tissues to regulate the localization, synthesis and degradation of hyaluronan which are essential to cells undergoing biological processes |
| P51884 | LUM | Lumican; Small leucine rich repeat proteoglycans; Belongs to the small leucine-rich proteoglycan (SLRP) family. SLRP class II subfamily |
| P18428 | LBP | Lipopolysaccharide-binding protein; Plays a role in the innate immune response. Binds to the lipid A moiety of bacterial lipopolysaccharides (LPS), a glycolipid present in the outer membrane of all Gram-negative bacteria. Acts as an affinity enhancer for CD14, facilitating its association with LPS. Promotes the release of cytokines in response to bacterial lipopolysaccharide; BPI fold containing |
| P15169 | CPN1 | Carboxypeptidase N catalytic chain; Protects the body from potent vasoactive and inflammatory peptides containing C-terminal Arg or Lys (such as kinins or anaphylatoxins) which are released into the circulation; M14 carboxypeptidases |
| P13796 | LCP1 | Plastin-2; Actin-binding protein. Plays a role in the activation of T-cells in response to costimulation through TCR/CD3 and CD2 or CD28. Modulates the cell surface expression of IL2RA/CD25 and CD69; EF-hand domain containing |
| P13671 | C6 | Complement component C6; Constituent of the membrane attack complex (MAC) that plays a key role in the innate and adaptive immune response by forming pores in the plasma membrane of target cells; Complement system |
| P12259 | F5 | Coagulation factor V; Central regulator of hemostasis. It serves as a critical cofactor for the prothrombinase activity of factor Xa that results in the activation of prothrombin to thrombin |
| P0DJI9 | SAA2 | Serum amyloid A-2 protein; Major acute phase reactant. Apolipoprotein of the HDL complex; Belongs to the SAA family |
| P0DJI8 | SAA1 | Serum amyloid A-1 protein; Major acute phase protein; Belongs to the SAA family |
| P05452 | CLEC3B | Tetranectin; Tetranectin binds to plasminogen and to isolated kringle 4. May be involved in the packaging of molecules destined for exocytosis; C-type lectin domain containing |
| P05155 | SERPING1 | Plasma protease C1 inhibitor; Activation of the C1 complex is under control of the C1- inhibitor. It forms a proteolytically inactive stoichiometric complex with the C1r or C1s proteases. May play a potentially crucial role in regulating important physiological pathways including complement activation, blood coagulation, fibrinolysis and the generation of kinins. Very efficient inhibitor of FXIIa. Inhibits chymotrypsin and kallikrein; Serpin peptidase inhibitors |
| P02768 | ALB | Serum albumin; Serum albumin, the main protein of plasma, has a good binding capacity for water, Ca(2+), Na(+), K(+), fatty acids, hormones, bilirubin and drugs. Its main function is the regulation of the colloidal osmotic pressure of blood. Major zinc transporter in plasma, typically binds about 80% of all plasma zinc; Belongs to the ALB/AFP/VDB family |
| P02750 | LRG1 | Leucine rich alpha-2-glycoprotein 1 |
| P02748 | C9 | Complement component C9; Constituent of the membrane attack complex (MAC) that plays a key role in the innate and adaptive immune response by forming pores in the plasma membrane of target cells. C9 is the pore- forming subunit of the MAC; Belongs to the complement C6/C7/C8/C9 family |
| P01011 | GIG25 | Serpin peptidase inhibitor, clade A (alpha-1 antiproteinase, antitrypsin), member 3; Although its physiological function is unclear, it can inhibit neutrophil cathepsin G and mast cell chymase, both of which can convert angiotensin-1 to the active angiotensin-2; Serpin peptidase inhibitors |
| P00751 | CFB | Complement factor B; Factor B which is part of the alternate pathway of the complement system is cleaved by factor D into 2 fragments: Ba and Bb. Bb, a serine protease, then combines with complement factor 3b to generate the C3 or C5 convertase. It has also been implicated in proliferation and differentiation of preactivated B- lymphocytes, rapid spreading of peripheral blood monocytes, stimulation of lymphocyte blastogenesis and lysis of erythrocytes. Ba inhibits the proliferation of preactivated B-lymphocytes; Belongs to the peptidase S1 family |
| P00738 | HP | Haptoglobin; As a result of hemolysis, hemoglobin is found to accumulate in the kidney and is secreted in the urine. Haptoglobin captures, and combines with free plasma hemoglobin to allow hepatic recycling of heme iron and to prevent kidney damage. Haptoglobin also acts as an Antimicrobial; Antioxidant, has antibacterial activity and plays a role in modulating many aspects of the acute phase response. Hemoglobin/haptoglobin complexes are rapidly cleared by the macrophage CD163 scavenger receptor expressed on the surface of liver Kupfer cells through an endocytic lysosomal degradation. |

**Supplementary Table 3:** List and taxonomic classifications of the PRS-related core microbial OTUs

| OTUs | Taxa annotation |
| --- | --- |
| OTU30210_Clostridiales<br>*spp* | k__Bacteria; p__Firmicutes; c__Clostridia; o__Clostridiales; f__; g__; s__ |
| OTU1090_Unassigned 2 | Unassigned |
| OTU174751_Clostridiu<br>*m spp* | k__Bacteria; p__Firmicutes; c__Clostridia; o__Clostridiales; f__Clostridiaceae; g__Clostridium; s__ |
| OTU175355_Ruminococ<br>*cus gnavus* | k__Bacteria; p__Firmicutes; c__Clostridia; o__Clostridiales; f__Lachnospiraceae; g__[Ruminococcus]; s__gnavus |
| OTU1954_Unassigned 1 | Unassigned |
| OTU198475_Ruminococ<br>*caceae spp 1* | k__Bacteria; p__Firmicutes; c__Clostridia; o__Clostridiales; f__Ruminococcaceae; g__; s__ |
| OTU212681_Blautia *spp* | k__Bacteria; p__Firmicutes; c__Clostridia; o__Clostridiales; f__Lachnospiraceae; g__Blautia; s__ |
| OTU27582_Klebsiella<br>*spp* | k__Bacteria; p__Proteobacteria; c__Gammaproteobacteria; o__Enterobacteriales; f__Enterobacteriaceae; g__Klebsiella; s__ |
| OTU30109_Streptococcu<br>*s spp 2* | k__Bacteria; p__Firmicutes; c__Bacilli; o__Lactobacillales; f__Streptococcaceae; g__Streptococcus; s__ |
| OTU358_Bacteroides<br>*ovatus 2* | k__Bacteria; p__Bacteroidetes; c__Bacteroidia; o__Bacteroidales; f__Bacteroidaceae; g__Bacteroides; s__ovatus |
| OTU3653_Bacteroides<br>*ovatus 1* | k__Bacteria; p__Bacteroidetes; c__Bacteroidia; o__Bacteroidales; f__Bacteroidaceae; g__Bacteroides; s__ovatus |
| OTU437_Streptococcus<br>*spp 1* | k__Bacteria; p__Firmicutes; c__Bacilli; o__Lactobacillales; f__Streptococcaceae; g__Streptococcus; s__ |
| OTU662_Streptococcus<br>*spp 3* | k__Bacteria; p__Firmicutes; c__Bacilli; o__Lactobacillales; f__Streptococcaceae; g__Streptococcus; s__ |
| OTU743_Unassigned 3 | Unassigned |
| OTU808_Lactobacillus<br>*spp* | k__Bacteria; p__Firmicutes; c__Bacilli; o__Lactobacillales; f__Lactobacillaceae; g__Lactobacillus; s__ |
| OTU87798_Bacteroides<br>*spp 2* | k__Bacteria; p__Bacteroidetes; c__Bacteroidia; o__Bacteroidales; f__Bacteroidaceae; g__Bacteroides; s__ |
| OTU87996_Bacteroides<br>*ovatus 3* | k__Bacteria; p__Bacteroidetes; c__Bacteroidia; o__Bacteroidales; f__Bacteroidaceae; g__Bacteroides; s__ovatus |
| OTU88234_Bacteroides<br>*spp 3* | k__Bacteria; p__Bacteroidetes; c__Bacteroidia; o__Bacteroidales; f__Bacteroidaceae; g__Bacteroides; s__ |
| OTU91647_Ruminococc<br>*aceae spp 2* | k__Bacteria; p__Firmicutes; c__Clostridia; o__Clostridiales; f__Ruminococcaceae; g__; s__ |
| OTU92227_Bacteroides<br>*spp 1* | k__Bacteria; p__Bacteroidetes; c__Bacteroidia; o__Bacteroidales; f__Bacteroidaceae; g__Bacteroides; s__ |

